# Polygenic Prediction of Cellular Immune Responses to Mumps Vaccine

**DOI:** 10.1101/2024.02.23.24303277

**Authors:** Brandon J. Coombes, Inna G. Ovsyannikova, Daniel J. Schaid, Nathaniel D. Warner, Gregory A. Poland, Richard B. Kennedy

## Abstract

In this report, we provide a follow-up analysis of a previously published genome-wide association study of host genetic variants associated with inter-individual variations in cellular immune responses to mumps vaccine. Here we report the results of a polygenic score (PGS) analysis showing how common variants can predict mumps vaccine response. We found higher PGS for IFNγ, IL-2, and TNFα were predictive of higher post-vaccine IFNγ (p-value = 2e-6), IL-2 (p = 2e-7), and TNFα (p = 0.004) levels, respectively. Control of immune responses after vaccination is complex and polygenic in nature. Our results suggest that the PGS-based approach enables better capture of the combined genetic effects that contribute to mumps vaccine-induced immunity, potentially offering a more comprehensive understanding than traditional single-variant GWAS. This approach will likely have broad utility in studying genetic control of immune responses to other vaccines and to infectious diseases.

## Introduction

Understanding mumps immunity is of paramount importance, especially considering the recent increase in mumps outbreaks despite widespread vaccination efforts^1^. The resurgence of mumps in the United States, even among highly immunized populations, underscores the need for comprehensive research to understand the driving factors behind vaccine-induced immunity such that better control this disease results. Our previous work, a genome-wide association study (GWAS) conducted by Ovsyannikova et al. in 2023, shed light on the host genetic variants associated with inter-individual variation in mumps vaccine-induced cellular immune responses^2^. This GWAS provided crucial insights into the genetic factors influencing mumps vaccine-induced immunity. Work done by our lab and others with mumps and other viral pathogens has demonstrated that vaccine-induced immune responses are multigenic^3,4^. Polygenic scores (PGS) summarize the estimated effects of many genetic variants on an individual’s phenotype, thereby offering a powerful approach to comprehensively assess the genetic basis of complex traits. PGS have been used to examine COVID-19 disease severity^5,6^ and risk of infection-induced sequelae^7^. The approach has also been proposed as an enrichment strategy to lower costs and increase success in clinical trials^8^. Surprisingly, PGS have never been applied to immunogenetic studies of vaccine responsiveness. We employed PGS to evaluate the genetic determinants underlying mumps vaccine-induced cellular immune responses in the same cohort previously studied by us^2^. This novel approach enables capture of combined genetic effects that contribute to mumps immunity, potentially offering a more comprehensive understanding than traditional single-variant GWAS. This approach holds the potential to significantly enhance our understanding of vaccine-induced immune responsiveness and could be used to reverse engineer new vaccine candidates based on a fuller understanding of immune response mechanisms^9^.

## Methods

The immune assays described herein are identical to those described in our previous vaccine studies^10-12^ and as used in our recent published GWAS^2^.

### Study Subjects

Our study cohort was a sample of 1,406 healthy children, older adolescents, and healthy adults selected from two cohorts with existing genotyping data from prior immunogenetics studies^10,11,13-15^. The demographic and clinical characteristics of these cohorts have been previously published^10,11,13,16^. Briefly, the cohort consisted of 1,406 healthy subjects (35.8 % females, 94.4 % European) with a mean time from last measles-mumps-rubella (MMR) vaccination to blood draw of 5.3 years (25 % and 75 % interquartile range/IQR 0.3, 15.5), and age at enrollment of 19.5 years (IQR 15.0, 23.0). The Institutional Review Boards of the Mayo Clinic (Rochester, MN) and the NHRC (Naval Health Research Center, San Diego, CA) approved the study, and written informed consent was obtained from each subject.

### Cytokine and Chemokine Response Measurements

We measured cytokine (IL-2, IL-6, IL-10, IFNα2a, IFNγ, IL-1β, and TNFα) and chemokine (IP-10, MCP-1, MIP1α, and MIP1β) responses from cultured PBMCs stimulated *in vitro* with mumps virus, as described and previously reported^12^. Here we focused on the five immune response outcomes for which we had previously found significant genome-wide SNP associations: IL-2, IFNγ, IL-1β, TNFα, and MCP-1. The coefficient of variation (CV) for these assays varied from 10% to 31% depending on the cytokine/chemokine.

### Genotyping and Imputation

The cohorts were genotyped using the Illumina Omni 1M array for school-age subjects recruited from the Rochester, MN area (n=964), and using the Illumina 550 and 650 SNP arrays (for the European and African-American subjects, respectively) for young adults recruited from the San Diego, CA area (n=807) ^10,11,14^. Each genotyping batch was filtered using standard quality control filters as described in Ovsyannikova et al. 2023 and then imputed using the TOPMed imputation reference panel^17^ retaining only well-imputed SNPs (dosage-R^2^ > 0.8) with MAF > 0.01.

Our initial GWAS of mumps-specific cytokines/chemokines was focused on the n=1,406 (903 males and 503 females) subjects with European ancestry (i.e., Rochester cohort, n=748 subjects, San Diego cohort, n=658 subjects). The covariates that were significantly associated with cytokine/chemokine levels were the following: age at last MMR vaccination; cohort (San Diego vs. Rochester); sex; age at blood draw; year of blood draw and the first 10 principal components (PCs) that were constructed from the genome wide SNP arrays. These associated covariates were used as independent variables in linear models with quantile-normalized cytokine/chemokine trait as dependent. The “residualized” cytokines from these models were then used as the dependent in each PGS association test.

### Polygenic Score (PGS) Analyses

We used a “leave-one-out” (LOO) approach to test whether the genetic prediction of cytokine response can predict observed cytokine response. In this LOO approach, GWASs were repeated as described in Ovsyannikova et al. (2023)^2^ except they were performed separately in the independent Rochester and San Diego cohorts using the same models. We then generated PGS within each cohort using the GWAS summary statistics from the other cohort. These LOO PGSs were constructed for each cytokine using PRSice2^18^. To account for linkage disequilibrium (LD) among SNPs, clumping was performed (clump-r2 0.1 clump-kb 250). We evaluated a series of p-value thresholds (*p*_*T*_ = 5e-7, 1e-7, 1e-6, 1e-5, 0.0001, 0.001, 0.01). Each PGS was then standardized. The PGS for each cytokine represents a genetically predicted cytokine. In each cohort, the genetically predicted cytokine (PGS) constructed with varying p-value thresholds was tested for association with the observed residualized cytokine using linear regression while adjusting for the first two principal components (PCs) of ancestry. The results from each LOO PGS analysis were then meta-analyzed across cohorts using the `meta R` package using a fixed-effect meta-analysis. In some cases, the PGS calculated under a stricter threshold (e.g., genome-wide significant variants only; p<5e-8) for one cohort was not possible to be estimated in the other cohort as no variants fit that threshold. In these cases, the results from only one cohort was reported for that p-value threshold.

## Results

### Study Sample Characteristics

The analyzed cohort (n=1,406) consisted of subjects with European (EUR, 94.4%) or Asian (5.6%) ancestries. The sex distribution was 903 males (64.2%) and 503 females (35.8%). The average age at enrollment was 19.5 years. Additional demographic details can be found in Ovsyannikova et. al.^2^ The current study focused on five immune outcomes: IL-2, IFNγ, IL-1β, TNFα, and MCP-1 (See Table 1) as these were the outcomes for which we had previously identified significant genome-wide SNP associations (IL-2, IFNγ, IL-1β, TNFα) or came close to the 5 × 10^−8^ threshold (MCP-1).

**Table 1.**
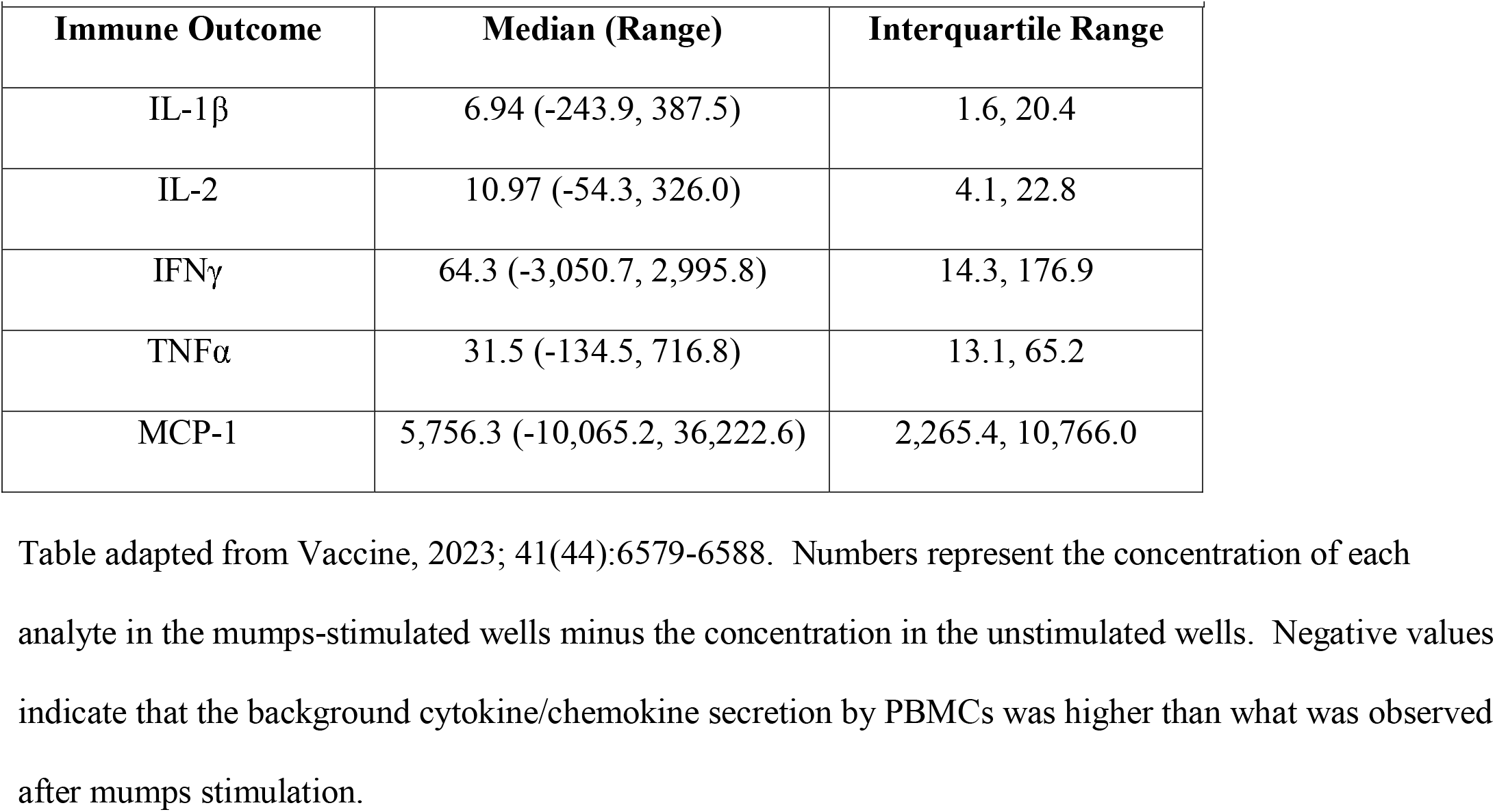
Cytokine and Chemokine Responses in the Study Cohort (n=1,406)

| Table 1. Cytokine and Chemokine Responses in the Study Cohort (n=1,406) |  |  |
| --- | --- | --- |
| Immune Outcome | Median (Range) | Interquartile Range |
| IL-1β | 6.94 (-243.9, 387.5) | 1.6, 20.4 |
| IL-2 | 10.97 (-54.3, 326.0) | 4.1, 22.8 |
| IFNγ | 64.3 (-3,050.7, 2,995.8) | 14.3, 176.9 |
| TNFα | 31.5 (-134.5, 716.8) | 13.1, 65.2 |
| MCP-1 | 5,756.3 (-10,065.2, 36,222.6) | 2,265.4, 10,766.0 |

Table adapted from Vaccine, 2023; 41(44):6579-6588. Numbers represent the concentration of each analyte in the mumps-stimulated wells minus the concentration in the unstimulated wells. Negative values indicate that the background cytokine/chemokine secretion by PBMCs was higher than what was observed after mumps stimulation.

### Polygenic Score Analysis Results with Mumps Cellular/Inflammatory Immune Responses

We first performed GWASs of vaccine outcomes (IFNγ, IL1β, IL-2, MCP1, and TNFα) separately in the cohort from Rochester (N = 728) and the cohort from San Diego (N = 599). In a LOO approach, we tested for association of LOO-PGS constructed within each of those cohorts using the oppositive cohort’s GWAS summary statistics and meta-analyzed the results from each cohort. Figure 1 shows the average variance explained by each LOO PGS for each cytokine at various p-value thresholds. In the meta-analysis, IFNγ (best *p*_*T*_ = 5e-8; *β*=0.16 (0.04); R^2^=3.1%; p-value = 9e-6), IL-2 (best *p*_*T*_ = 1e-5; *β*=0.13 (0.03); R^2^=2.1%; p-value = 2e-7), and TNFα (best *p*_*T*_ = 1e-6; *β*=0.11 (0.02); R^2^=1.2%; p-value = 0.004) were each significantly predicted by their respective PGS.

**Figure 1.**
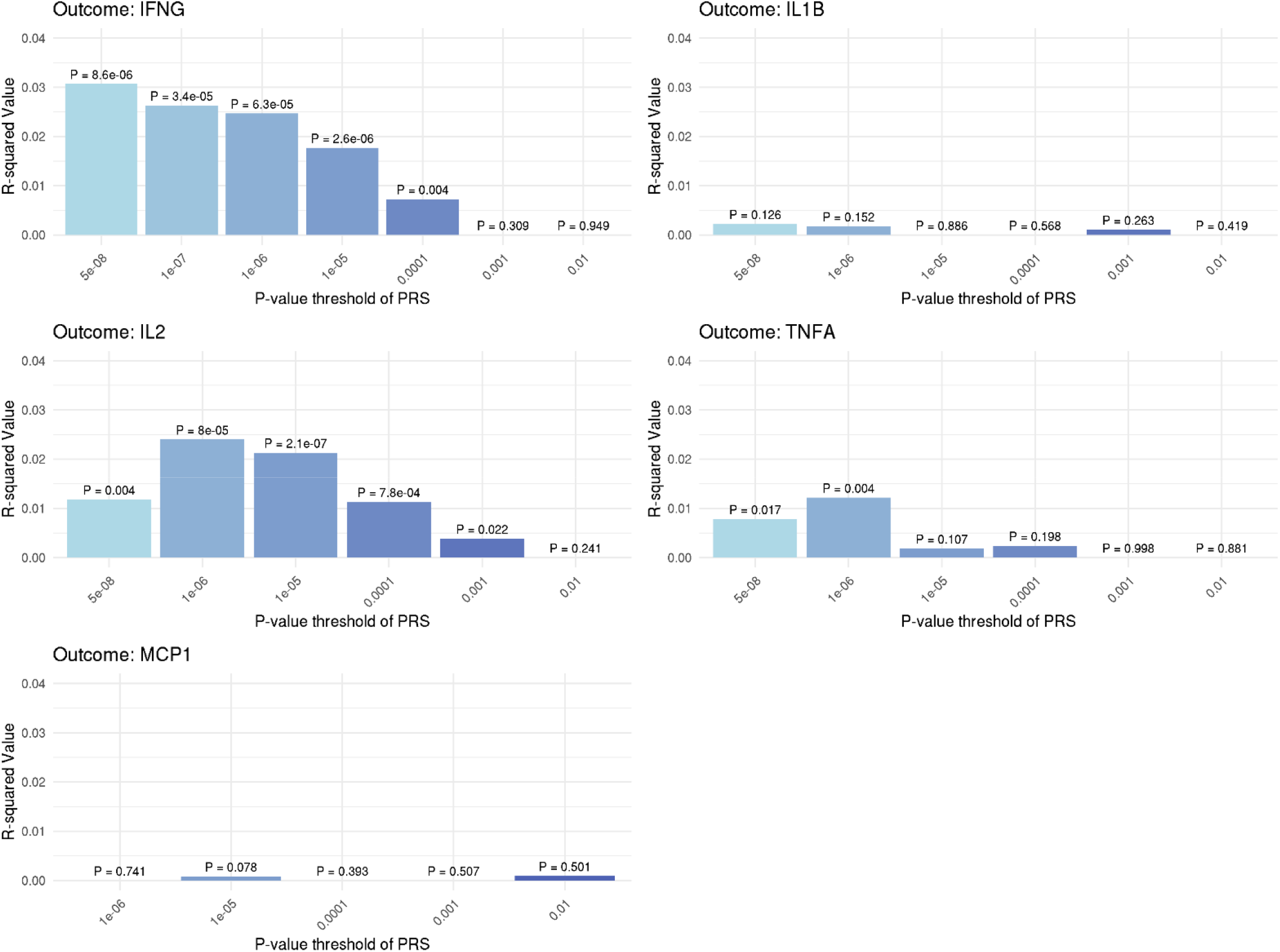
Bar plots depicting the average variance explained in each residualized cytokine outcome across cohorts when using a LOO PGS for each cytokine with different p-value thresholds of inclusion of variants. The meta-analysis p-value for association is annotated above each bar.

We noted weak signals for IL1β and MCP-1, suggesting that the genetic contributions of the secretion of these factors is weaker than what is observed for the other factors. For IL-2 and TNFα, lowering the p-value threshold to include more SNPs in the analysis resulted in both an increase in variance explained and a lower meta-analysis p-value. This increased signal tapered off as the p-value threshold for including SNPs in the PRS increased to 0.0001. This suggests that there are a limited number of additional genetic factors influencing secretion of these factors (i.e., control of these cytokines is likely multi-genic but not poly-genic). For IFNγ, the genome-wide significance p-value threshold explained the most variation. However, the PGS was not calculated at this threshold (nor the p<1e-7 and 1e-6 thresholds) in the Rochester cohort because in the San Diego cohort no variant met these thresholds. Therefore, the strictest p-value threshold (p<1e-5) with available variants in both cohorts actually achieved the smallest meta-analysis p-value.

Our initial results suggested there are some SNPs below the genome-wide significance level that appear to improve prediction, therefore we examined the SNPs reaching the 1 × 10^−6^ threshold and looked up the gene most connected to each SNP according to the variant-to-gene (V2G) score (Table 2)^19,20^. For IFNγ, 6 additional variants (mostly in the HLA region) were identified as potentially contributing to the variance explained. For IL-2, 7 additional variants in the HLA region were identified. For TNFa, we identified an additional SIGLEC 5/14 variant as well as a variant in NCOR2 and a variant in ATXN7L3B.

**Table 2.** SNPs Associated with p < 1e-6 with Increased Outcome Variance Explained by PRS.

| Chr | BP | A1/A2 | SNP | p-value | Outcome | Nearest gene | Best evidenced gene (V2G) |
| --- | --- | --- | --- | --- | --- | --- | --- |
| 6 | 32443876 | C/T | rs28895246 | 5.6e-13 | IFNγ | HLA region | n/a |
| 6 | 32190620 | T/C | rs3134931 | 7.8e-08 | IFNγ | NOTCH4 | HLA-DRB5 |
| 2 | 204450276 | T/C | rs12693990 | 6.4e-07 | IFNγ | RAPH1 | FAM117B |
| 6 | 31097814 | C/T | rs6906565 | 1.6e-07 | IFNγ | PSORS1C2 | HLA-B, HLA-C, CCHCR1, PSORS1C1 |
| 6 | 31461613 | T/C | rs2252937 | 1.0e-07 | IFNγ | MICB | HLA-C |
| 6 | 32586236 | C/A | rs11751024 | 2.1e-07 | IFNγ | HLA region | n/a |
| 6 | 32230354 | A/G | rs3132972 | 1.6e-10 | IL-2 | NOTCH4 | HLA-DRB5 |
| 6 | 32428772 | A/G | rs9268839 | 1.1e-16 | IL-2 | HLA-DRA | HLA-DQA2 |
| 6 | 32678867 | G/A | rs4476871 | 2.1e-12 | IL-2 | HLA-DQA2 | HLA-DQA2 |
| 6 | 31101849 | A/G | rs9468872 | 3.3e-07 | IL-2 | PSORS1C2 | HLA-C, CCHCR1, PSORS1C1 |
| 6 | 32428397 | C/A | rs17202787 | 7.1e-07 | IL-2 | HLA-DRA | HLA-DRB5 |
| 6 | 32586236 | C/A | rs11751024 | 2.0e-07 | IL-2 | HLA region | n/a |
| 6 | 32684468 | C/T | rs5024432 | 5.9e-07 | IL-2 | HLA-DQA2 | HLA-DRB5 |
| 19 | 52130638 | C/A | rs872629 | 8.2e-13 | TNFα | SIGLEC5 | SIGLEC14 |
| 12 | 75052038 | A/G | rs7302396 | 1.1e-07 | TNFα | ATXN7L3B | ATXN7L3B |
| 12 | 125160661 | G/A | rs838502 | 4.8e-07 | TNFα | NCOR2 | NCOR2 |

BP coordinates = genomic location; Nearest gene to SNP based on canonical distance; V2G score is a weighted measure developed by Open Targets Genetics.

## Discussion

In our cohort of 1,406 mumps vaccine recipients, we found that higher polygenic scores for IFNγ, IL-2, and TNFα were all predictive of higher post-vaccination cytokine responses to *in vitro* stimulation with inactivated mumps virus. The *in vitro* stimulation elicited a recall response from virus-specific T cells without the complications of viral infection, acting as a probe allowing us to evaluate the magnitude and quality of the T cell response that would be expected upon exposure to mumps virus. Furthermore, the PRS clearly indicates that including more variants than those that were genome-wide significant improved the strength of association. While widely used in some fields (e.g., psychiatry), our study demonstrates that PGS scores can also be calculated for vaccine immune response and that such scores can be associated with the same immune outcome in independent cohorts. The increased variation in outcome explained by the PRS indicate that there is additional power to including information from variants not reaching the standard genome-wide threshold. Thus, PRS approaches can supplement GWAS by providing additional information about genetic control of viral vaccine responses.

We have already reported genetic associations with HLA loci and mumps-specific IFNγ and IL-2 secretion^2^. Our PRS found additional polymorphisms in the same region (HLA) also contributed to the variation in IL-2 secretion; however, for IFNγ we identified not only additional HLA loci, but also SNPs near *FAM117B* and *RAPH1* genes that may potentially influence cytokine secretion. *FAM117B* (also known as *ALS2CR13)*, is a gene associated with the neurodegenerative disorders juvenile ALS (amyotrophic lateral sclerosis) and mast syndrome.^21^ RAPH1 encodes for an adapter protein that functions in cell migration. Interestingly, this gene is also associated with juvenile ALS. With respect to TNFα, our PRS identified one additional SNP in SIGLEC5/14 as well as a SNP in *ATXN7L3B* and a SNP in *NCOR2. ATXN7L3B* encodes for the Ataxin 7 like protein 3B, which regulates histone deubiquitination, while *NCOR* encodes for a nuclear receptor co-repressor that interacts with retinoic acid receptor. Immune-related functions for these genes have not been identified and further investigation will be necessary to determine what role they might play in the cytokine response to mumps vaccination.

To the best of our knowledge, this study represents the first use of use of polygenic scores to study genetic influences on cellular immune responses after vaccination. Our results indicate that this approach has promise, as it identified additional genetic variants potentially contributing to inter-individual variation in immune responses to mumps virus stimulation. These additional variants/regions may provide information regarding the genes/pathways involved in regulating the immune response and represent targets for functional studies to elucidate the biological mechanisms behind the statistical associations. It is likely that calculation of polygenic risk scores will complement existing GWAS approaches when investigating multigenic control of immune responses.

## Data Availability

All data produced in the present work are contained in the manuscript.

## Acknowledgments

We thank the Mayo Clinic Vaccine Research Group staff and the study participants.

